# Post-lockdown Dynamics of COVID-19 in New York, Florida, Arizona, and Wisconsin

**DOI:** 10.1101/2020.12.28.20248967

**Authors:** Sherry Scott, Keisha J. Cook, Kamal Barley

## Abstract

The COVID-19 pandemic is widely studied as it continues to threaten many populations of people especially in the USA, the leading country in terms of both deaths and cases. Although vaccines are being distributed, control and mitigation strategies must still be properly enforced. More and more reports show that the spread of COVID-19 involves infected individuals first passing through a pre-symptomatic infectious stage in addition to the incubation period and that many of the infectious individuals are asymptomatic. In this study, we design and use a mathematical model to primarily address the question of who are the main drivers of COVID-19 - the symptomatic infectious or the pre-symptomatic and asymptomatic infectious in the states of Florida, Arizona, New York, Wisconsin and the entire United States. We emphasize the benefit of lockdown by showing that for all four states, earlier and later lockdown dates decrease the number of cumulative deaths. This benefit of lockdown is also evidenced by the decrease in the infectious cases for Arizona and the entire US when lockdown is implemented earlier. When comparing the influence of the symptomatic infectious versus the pre-sympomatic/asymptomatic infectious, it is shown that, in general, the larger contribution comes from the latter group. This is seen from several perspectives, as follows: (1) in terms of daily cases, (2) in terms of daily cases when the influence of one group is targeted over the other by setting the effective contact rate(s) for the non-targeted group to zero, and (3) in terms of cumulative cases and deaths for the US and Arizona when the influence of one group is targeted over the other by setting the effective contact rate(s) for the non-targeted group to zero. The consequences of the difference in the contributions of the two infectious groups is simulated in terms of testing and these simulations show that an increase in testing and isolating for the pre-symptomatic and asymptomatic infectious group has more impact than an increase in testing for the symptomatic infectious. For example, for the entire US, a 50% increase in testing for the pre-symptomatic and asymptomatic infectious group results in a 25% decrease in deaths as opposed to a lower 6% decrease in deaths when a 50% increase in testing rate for the symptomatic infectious is implemented. We also see that if the testing for infectious symptomatic is kept at the baseline value and the testing for the pre-symptomatic and asymptomatic is increased from 0.2 to 0.25, then the control reproduction number falls below 1. On the other hand, to get even close to such a result when keeping the pre-symptomatic and asymptomatic at baseline fitted values, the symptomatic infectious testing rate must be increased considerably more - from 0.4 to 1.7. Lastly, we use our model to simulate an implementation of a natural herd immunity strategy for the entire U.S. and for the state of Wisconsin (the most recent epicenter) and we find that such a strategy requires a significant number of deaths and as such is questionable in terms of success. We conclude with a brief summary of our results and some implications regarding COVID-19 control and mitigation strategies.

## 1. Introduction

The coronavirus, known as COVID-19, emerged in Wuhan, China in December 2019, and has since then spread to all countries on earth. As of 12/10/2020, the United States has reported 15,535,565 cases and 291,403 deaths and although the leading country in both deaths and cases, the U.S. did not implement a national pandemic control initiative. Instead, many states and cities implemented their own initiatives at various times, and the transmission of COVID-19 has varied from state to state. States also implemented their own testing initiatives. Around 12-10-2020, the reported testing rates are as follows: Arizona testing at a rate of 347.7 tests per 100,000 people, Florida testing at a rate of 497.2 tests per 100,000 people, New York testing at a rate of 985.7 tests per 100,000 people, and Wisconsin testing at a rate of 562.4 tests per 100,000 people [2]. With regard to mask usage, the observed mask usage percentage in the US has increased since the beginning of the pandemic. From March 1, 2020 to May 1, 2020, the mask use percentage increased from 5% to 40%. From May 1, 2020 to November 22, 2020, the mask use percentage increased from 71%. Social distancing has also played a role in the transmission of COVID-19. From March 8, 2020 to April 4, 2020, the US experienced a 53% decrease in mobility. From April 4, 2020 to June 24, 2020, the decrease in mobility changed to only −22%. From June 24, 2020 to November 22, 2020, the mobility oscillated between −18% and −25% [3]. The social distancing statistics are due to the fact that different states implemented stay-at-home periods for various intervals of time. All of these factors play a part in the transmission of COVID-19 in the US as a whole, and effectively in each state. Although we do not consider all of these measures directly, we do discuss the effect that lockdown has on the number of cases and deaths. We do include mask usage, and we use a different mask compliance value for each period per state [14]. To study social distancing, we fit the values for the effective contact rate for each time period per state and per time period.

Mathematical modelling can help us better understand how COVID-19 cases and mortality is affected based on when and how long lockdown periods are enforced. Modelling can also help us identify the main drivers of the disease and give insight on how best to use the standard control and mitigation strategies. We expand on the standard SIR model, which uses a system of ordinary differential equations to model disease spread through multiple compartments over time. In this study, we do not consider a time-varying system.

Prior studies consider susceptible individuals transitioning to exposed and then to infectious. We include a pre-symptomatic compartment for individuals who are infectious before the onset of symptoms. We also consider individuals who were tested, resulting in an asymptomatic or symptomatic classification, as well as the population of people who may have self-isolated after being exposed by an infected individual. It is known that over the course of the pandemic, hospitals have been highly stressed and over-crowded due to the influx of COVID-19 patients. In order to study mortality due to the pandemic, we also include compartments that keep track of the number of patients admitted to the hospital and the ICU.

We explore scenarios where lockdown is started earlier or extended past the reported date. We show that both scenarios decrease the number of deaths. We can use these type of simulations to predict future mortality populations and use the information to help with determining when lockdown periods should start and how long the lockdown period should be enforced.

Finally, in response to the increase in public discussions of a natural herd immunity approach, we use our model to run some basic scenarios for implementing such a strategy in the state of Wisconsin and the US as a whole. We simulate the number of deaths that would occur before herd immunity is achieved.

## 2. Main Questions

The overall purpose of this study is to explore the post-lockdown dynamics of COVID-19 and in the process, explore the notion that the main drivers of COVID19 are the asymptomatic and pre-symptomatic infectious individuals. Because both the asymptomatic and pre-symptomatic infectious spread COVID-19 without showing symptoms we group these two compartments together and compare that group to the symptomatic infectious. More specifically, we use a mathematical model to address the following questions for New York, Arizona, Florida, Wisconsin, and in some cases, the entire US. The section of the paper in which each question is addressed is given after the question.

- How do changes in lockdown dates (i.e., earlier or later lockdown dates) affect the number of deaths and cases? (Section 4.3.1)
- How does the contribution of the pre-symptomatic and asymptomatic infectious individuals compare to that of the symptomatic infectious in terms of cases and deaths? (Section 4.3.2)
- If we isolate all symptomatic infectious individuals or all asymptomatic and pre-symptomatic infectious individuals, how will this impact deaths and cases? (Section 4.3.3)
- How might a minimum testing rate for pre-symptomatic and asymptomatic compare to that for the symptomatic infectious individuals in order to obtain a reproduction number below 1? (section 4.3.4)
- How many deaths will occur in a natural herd immunity simulation for Wisconsin and the entire US? (Section 4.4)

### 2.1. COVID-19

In order to assess and deliver guidelines for individuals to avoid coronavirus infection, the CDC has implemented models equipped with the many stages of the virus [1]. A person who has been infected is considered pre-symptomatic (an individual who is infectious but did not show symptoms at the time of testing) or asymptomatic (a person who is infectious and does not show symptoms throughout the course of the infection). The CDC built pandemic planning scenarios varying the infectiousness of asymptomatic individuals compared to symptomatic, varying the percent of asymptomatic individuals, and varying the percentage of transmission of pre-symptomatic individuals. In this paper, we use similar methods to build our model and determine parameter values for the locations and time periods of interest. Figure 1 shows the stages of COVID-19, especially the incubation period, infectious period, pre-symptomatic period, and symptomatic period. It should be noted that a pre-symptomatic infectious individual can become either asymptomatic or symptomatic infectious.

**Figure 1:**
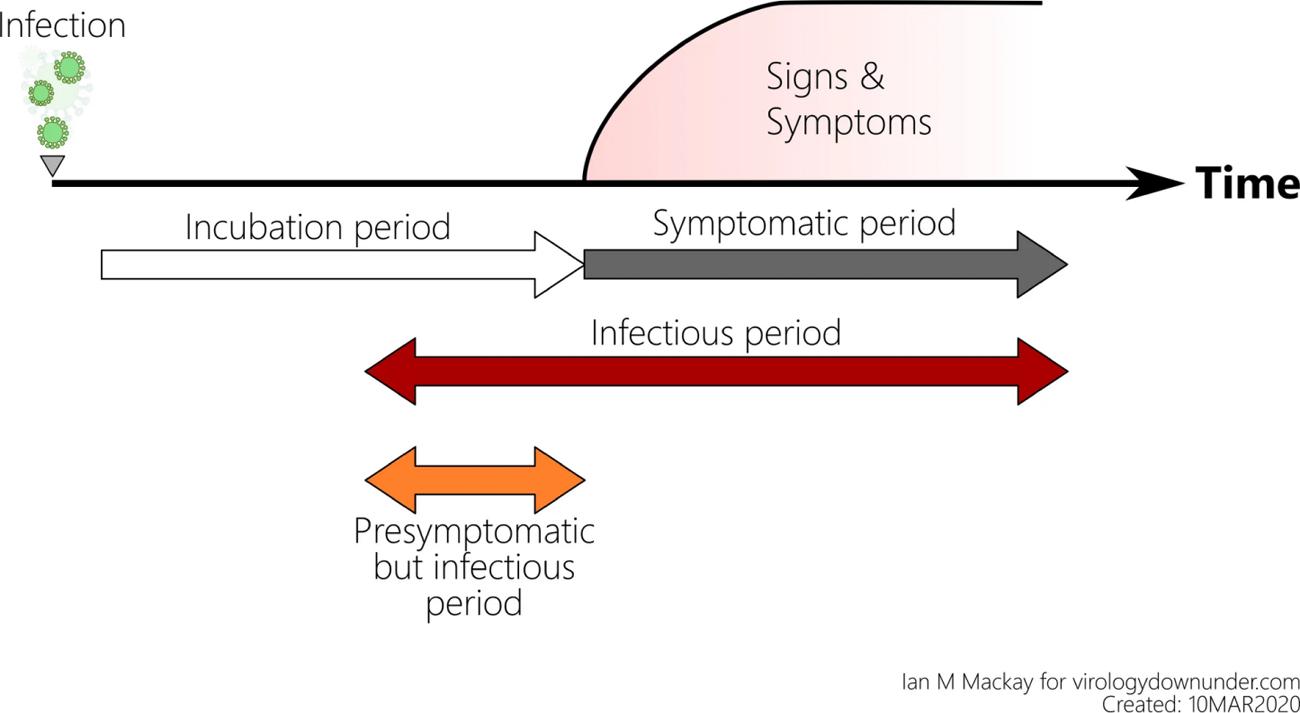
The Stages of COVID-19

**Figure 2:**
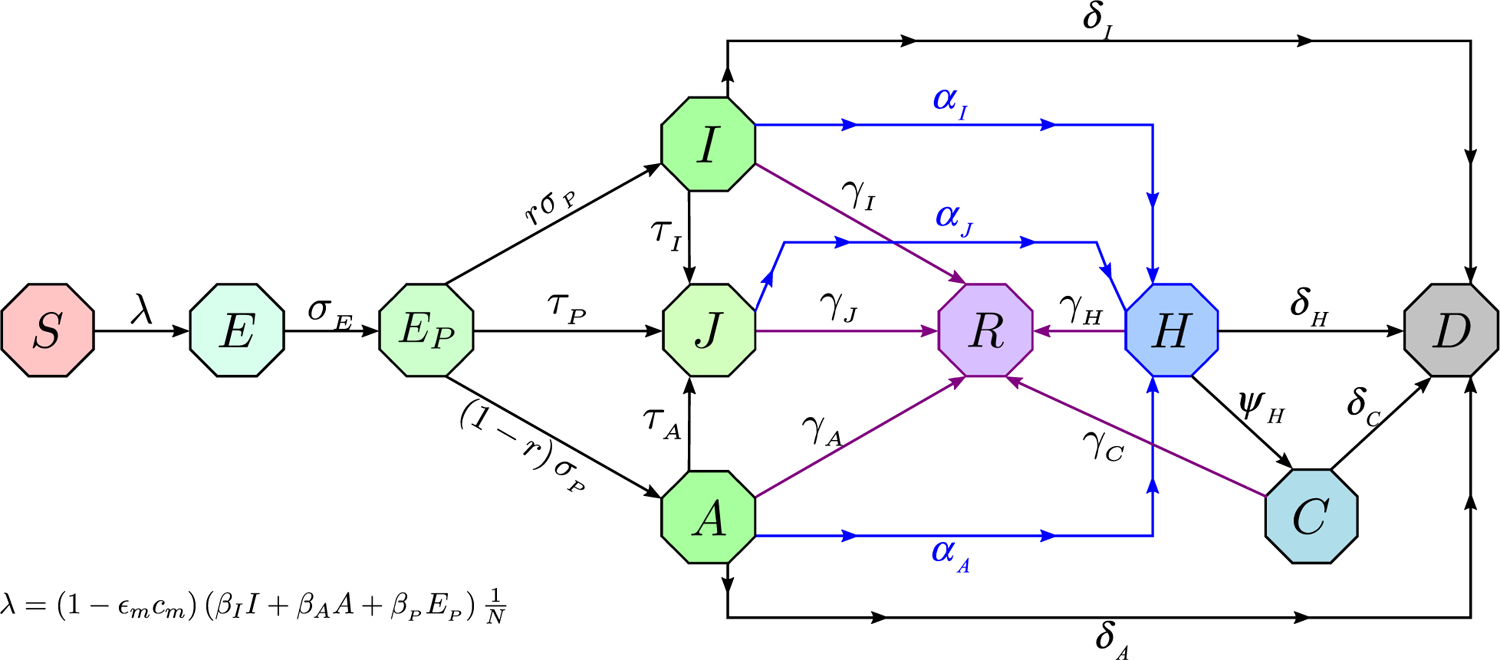
Schematic flow diagram of the model

We are interested in modeling COVID-19 in the United States as a whole, as well as focusing on four states. We consider New York (Northeast), which started as an epicenter in the US. We consider Florida (Southeast) and Arizona (Southwest) due to the fact that they became hot spots with significant numbers of daily cases and overwhelming numbers in their healthcare facilities. Lastly, we consider Wisconsin (Midwest), which became a hot spot later in the year. Table 1 shows the differences between the states with regards to the number of cumulative deaths.

**Table 1:**
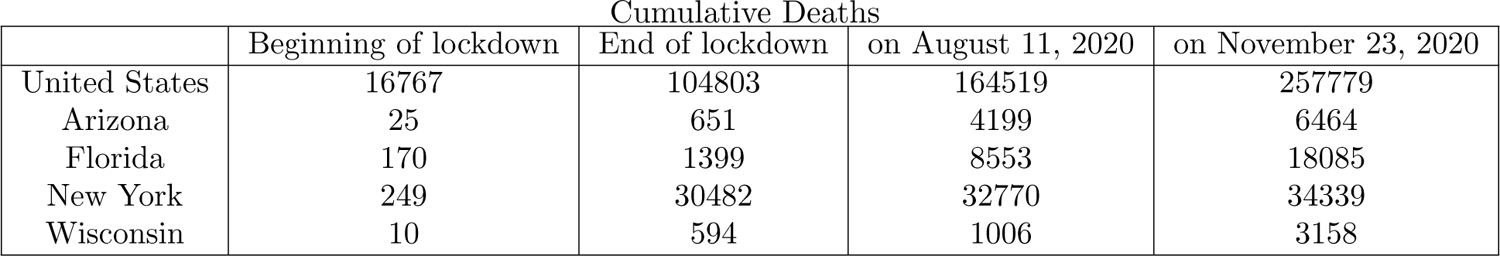
This table gives the cumulative deaths for each state and the United States. The lockdown dates depend on the state.

## 3. Methods

### 3.1. Model with pre-symptomatic compartment and face masks

We use a modified Kermack-McKendrik-type epidemic (no human demography) model to better understand the dynamics of COVID-19. Since a main objective of this study is to analyze the role of infectious individuals who show no symptoms, the model (2) includes a pre-symptomatic compartment as the foundation for parameter estimation. We use a deterministic susceptible, exposed, pre-symptomatic, symptomatically-infectious, asymptomatically-infectious, self-isolated, hospitalized, recovered, and ICU patients modeling framework, with the classes denoted as *S*(*t*), *E*(*t*), *E_P_* (*t*), *I*(*t*), *A*(*t*), *J*(*t*), *H*(*t*), *R*(*t*), *C*(*t*) respectively; we also include *D*(*t*) to track deaths. The model also includes the mitigation/control interventions of face mask usage (with a compliance parameter as well as a face mask efficacy parameter) and testing/detection implementation for each of the infectious classes.

### 3.2. Data Collection

We obtain the observed cumulative deaths and cases data for the states of Arizona, Florida, New York and Wisconsin from the COVID-19 Data Repository by the Center for Systems Science and Engineering (CSSE) at Johns Hopkins University (2020). The repository contains data beginning at January 22, 2020 (the marked beginning of the pandemic in the US), however, we focus primarily on the data from March 1, 2020 through October 13, 2020. Using these data sets we determine the initial conditions for the number of cases and deaths for a given time period.

The model has 24 parameters and we use values from the literature for 18 of these and estimate the remaining 6 by fitting the model to the observed cumulative mortality data for each state. The parameters that are estimated are as follows: the effective contact rate for the symptomatically-infectious individuals *β_I_*, the effective contact rate for the asymptomatically-infectious individuals *β_A_*, the effective contact rate for pre-symptomatically-infectious individuals *β_P_*, the rate at which presympotomatically-infectious individuals self-isolate *τ_P_*, the rate at which asympotomatically-infectious individuals self-isolate *τ_A_*, and the hospitalization rate for self-isolating individuals *α_J_*. Parameter fitting was performed using a non-linear sum-of-squares estimate - i.e. determining the best parameters set that minimizes the sum of the square of the difference between the model outputs for death and the observed values for deaths.

Our model for the transmission dynamics of COVID-19 is given by the following deterministic system of non-linear differential equations.

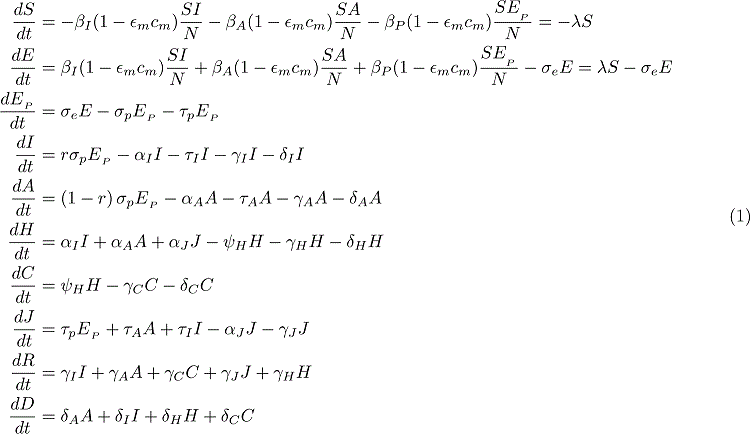

where *N* (*t*) = *S* + *E* + *EP* + *I* + *A* + *H* + *J* + *C* + *R* is the total population at time *t*. Let *E_m_* be the efficacy of face masks to prevent transmission and acquisition of infection, *c_m_* the compliance in face mask usage in the community (0 *< c_m_ ≤* 1), *σ* the progression rate from exposed (*E*) to infectious classes (*I* or *A*), *r* the fraction of exposed individuals who show clinical symptoms at the end of the incubation period, *τ_I_* the rate self-quarantined symptomatically-infectious humans self-isolate, *γ_I_*, *γ_A_*, *γ_J_*, *γ_H_*, *γ_C_* the recovery rates for the subscripted population, *ψ_H_* the rate of ICU admission for hospitalized individuals, *α_I_* the hospitalization rate for symptomatically-infectious individuals, *α_A_* the hospitalization rate for asymptomatically-infectious individuals, *δ_I_* the disease-induced mortality rate for symptomatically-infectious individuals, *δ_A_* the disease-induced mortality rate for asymptomatically-infectious individuals, *δ_H_* the disease-induced mortality rate for hospitalized individuals, *δ_C_* the disease-induced mortality rate individuals in ICU. We assume that hospitalized individuals do not come in contact with the general population.

## 4. Results

### 4.1. Computation of Reproduction Numbers

The basic reproduction number *R*_0_ is the average number of secondary infections produced when one infected individual is introduced into a host population of susceptible individuals. For our model’s *R*_0_, we have *R*_0_ = *R*_0_*_A_* + *R*_0_*_I_* + *R*_0_*_P_* where

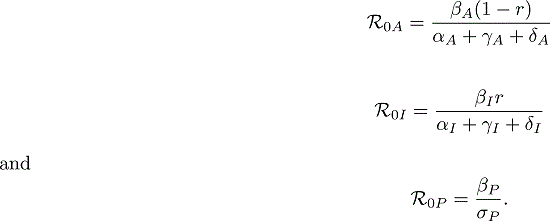

The control reproduction number *R_C_* is the average number of new cases generated by a typical infectious individual introduced into a host population of susceptible individuals with some control measures/interventions in place. Using the next generation operator method and notation found in [17] we have the following computation for the control reproduction number *R_C_*. If we take the column vector (*E E_P_ I A*) representing the compartments of infected, we have the associated next generation matrices, *F* and *V*, for the new infection terms and the transition terms are given respectively as

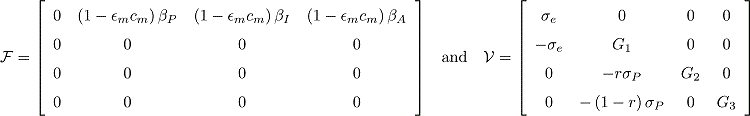

where, *G*_1_ = *σ_P_* + *τ_P_*, *G*_2_ = *α_I_* + *γ_I_* + *δ_I_* + *τ_I_*, *G*_3_ = *α_A_* + *γ_A_* + *δ_A_* + *τ_A_*. *R_C_* is the spectral radius of the next generation matrix given by,

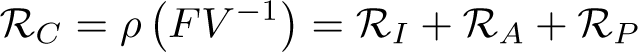

where 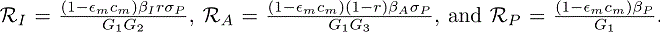. In absence of interventions, i.e. with *τ_I_* = *τ_A_* = *τ_P_*= *τ_A_*= *E_m_*= *c_m_*= 0, we get the basic reproduction number for model (1), *R*_0_, as given earlier.

**Theorem 1.** The disease-free equilibrium (DFE) of the model 1 is locally-asymptotically stable if R_C_ < 1*, and unstable if R_C_ >* 1.

A basic implication of Theorem 1 is that in order to control the COVID-19 outbreak (to not generate more), it suffices to keep *R_C_ <* 1.

### 4.2. Goodness of Fit

We analyze how well our model fits the data from John Hopkins using the T-Test in *R*. The T-Test measures the difference between two sets of continuous data. We want to compare two sample means to determine if there is a statistically significant difference between the model and the data. We use a paired T-test where each vector has the same amount of entries and the values are taken from the same independent variable. The T-Test assumes that the data in each vector is normally distributed and that they have approximately equal variances. To test for normality, we use the Shapiro-Wilk Test in *R*. The null hypothesis for this test is that the values in the vector are normally distributed. If the p-value is greater than 0.05, then the distribution of the values in the vector are not significantly different from the normal distribution. I.e. we fail to reject the null hypothesis. We can visualize this using a Q-Q plot. We assume normality if all the points fall along the reference line. We then check the variances of the values in the two vectors using the standard variance equation from probability. Let the null hypothesis for the T-Test be that the difference between the two groups is 0. The T-statistic is defined as

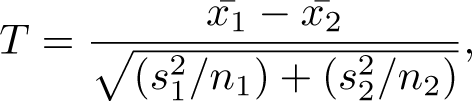

where, *x*^-^_1_ is the mean of the data, *x*^-^*_s_* is the mean of the model values, *s*^2^ is the variance of the data, *s*^2^ is the variance of the model values, *n*_1_ is the sample size of the data, and *n*_2_ is the sample size of the model values. We conclude that the model for each state, cumulative death population, during the lockdown period does indeed fit the data. Table 2 shows the resulting p-values of the T-Test for each state and the United States as a whole during the lockdown period. The goodness of fit plots can be found in the appendix (section 6.3) in Figures 17, 18, 19, 20, and 21.

**Table 2:** Goodness of Fit for the T-Test. This table gives the p-values and T-statistic for each test for each state and the United States during the lockdown period. The daily column denotes the model vs the data of the daily deaths. The cumulative column denotes the model vs the data of the cumulative deaths.

|  | Daily p-value | Daily T-statistic | Cumulative p-value | Cumulative T-statistic |
| --- | --- | --- | --- | --- |
| United States | 0.9722 | 0.035037 | 0.7839 | 0.27581 |
| Florida | 0.9083 | 0.11621 | 0.4947 | 0.69165 |
| Arizona | 0.6002 | -0.52796 | 0.0004622 | 3.7845 |
| New York | 0.8781 | -0.15396 | 0.007702 | 2.749 |
| Wisconsin | 0.02353 | -2.323 | 2.2e-16 | -17.953 |

### 4.3. Simulated Epidemics

We fit our model using the cumulative and daily mortality data for Florida, Arizona, Wisconsin, and New York over the pre-lockdown, lockdown, and post-lockdown periods based on the state’s respective decisions. Florida, Arizona, New York, and Wisconsin pre-lockdown period is defined from March 10, 2020 to April 3, 2020, March 21, 2020 to March 31, 2020, March 2, 2020 to March 22, 2020, and March 19, 2020 to March 25, 2020 respectively. The lockdown period is defined from April 3, 2020 to May 4, 2020, March 31, 2020 to May 15, 2020, March 22, 2020 to May 28, 2020, March 25, 2020 to May 26, 2020 respectively. For the entire US the pre-lockdown and post-lockdown dates are January 22, 2020 to April 7, 2020 and April 7, 2020 to May 28, 2020, as given in [14]. The post-lockdown periods begin at the end of the prescribed lockdown period for each state and for most of our results ends on August 11, 2020. Figure 3 shows the fit for our model with the cumulative deaths data during the lockdown period.

**Figure 3:**
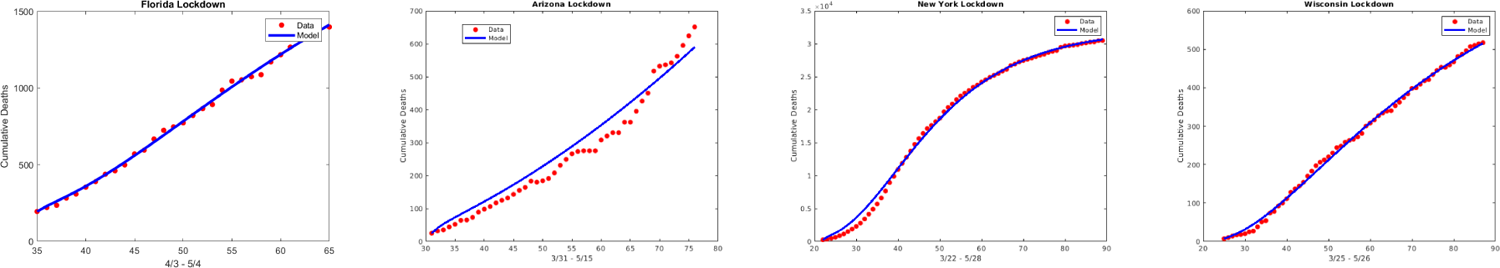
Data fitting of the model using COVID-19 mortality data for the states of Florida, Arizona, New York and Wisconsin during their respective lockdown periods. The plots show the model (blue) against the data (red).

The state variables and the parameters for our model are given in the Appendix in Tables 3 and 4 respectively.

#### 4.3.1. Lockdown starting earlier or later

For all of the states, the first lockdown (and for most states, the only lockdown so far) was implemented in the month of March. Since an earlier lockdown would have most likely prevented the disease from gaining a foothold in these areas, when we simulate an earlier lockdown, we find that the number of cumulative deaths decreases significantly for each state. This decrease occurs each time we add one week to the beginning of the lockdown period - see Figure 4.

**Figure 4:**
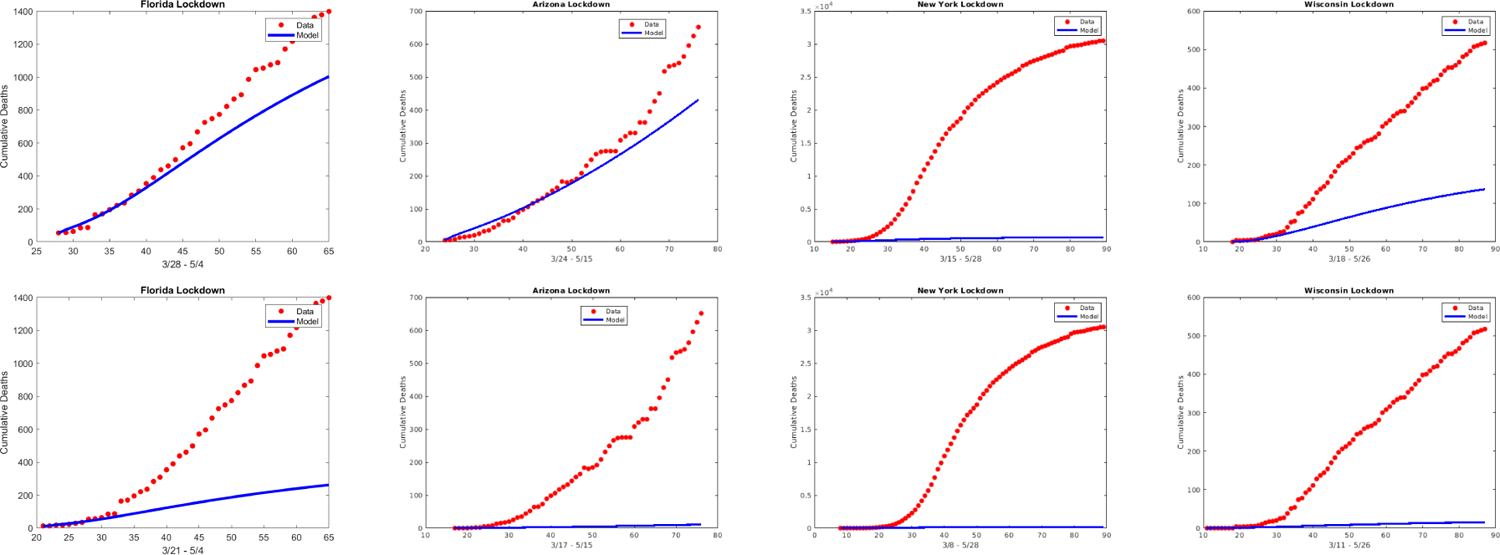
**Row 1**: The cumulative deaths for each state if lockdown began 1 week earlier. **Row 2**: The cumulative deaths for each state if lockdown began 2 weeks earlier.

In general, an earlier lockdown also results in a decrease in daily cases for both the symptomatic and asymptomatic/pre-symptomatic compartments, as shown in the daily cases plots for Arizona and the entire US in Figure 5.

**Figure 5:**
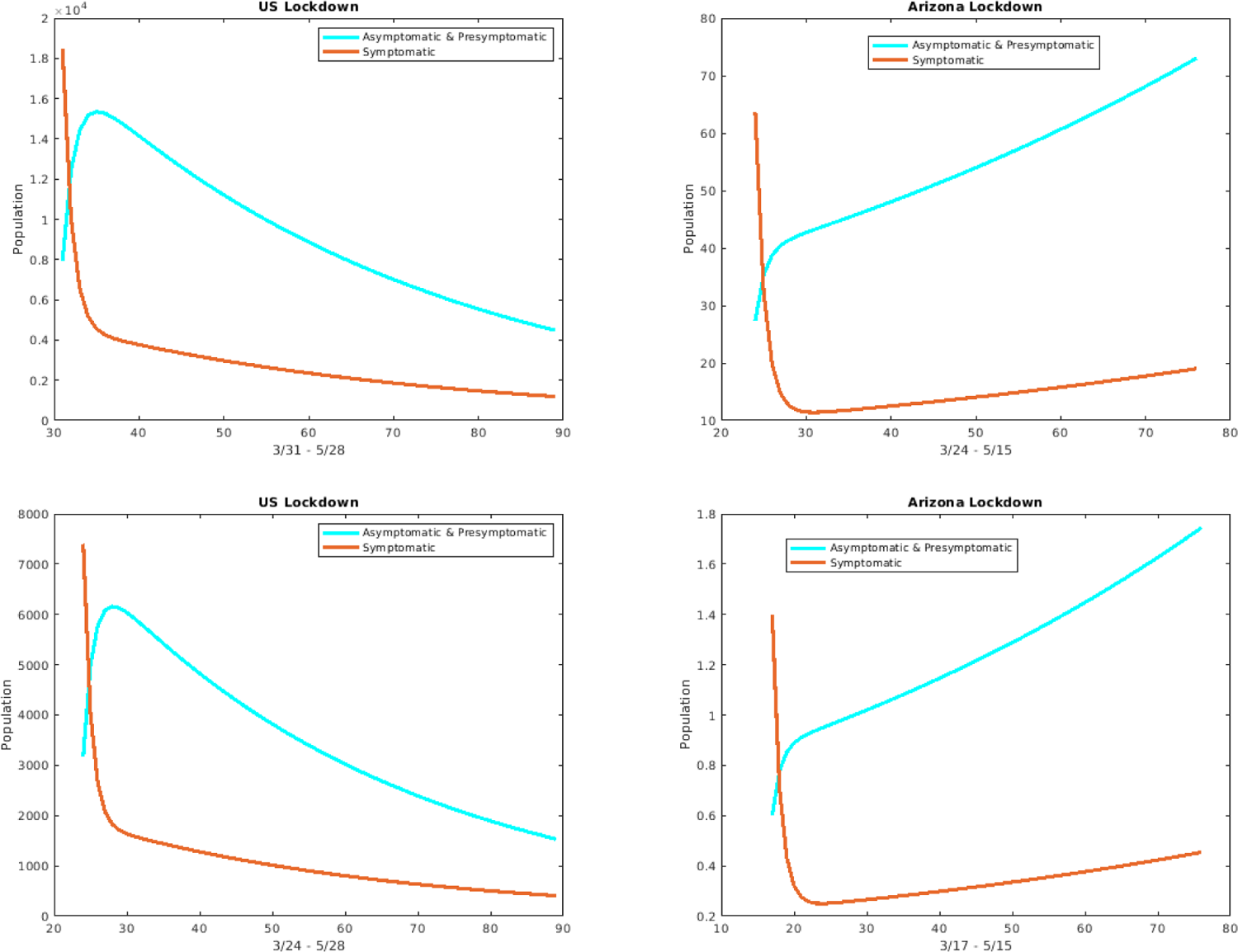
Daily cases for the asymptomatic/pre-symptomatic individuals (cyan curve) versus the symptomatic individuals (red curve) for the entire US and Arizona. **Row 1**: If lockdown began 1 week earlier. **Row 2**: If lockdown began 2 weeks earlier.

On the other hand, if we use the original lockdown start dates but extend the lockdown periods by 2 and 4 weeks for each state, we again see a decrease in cumulative deaths as shown in Figure 6.

**Figure 6:**
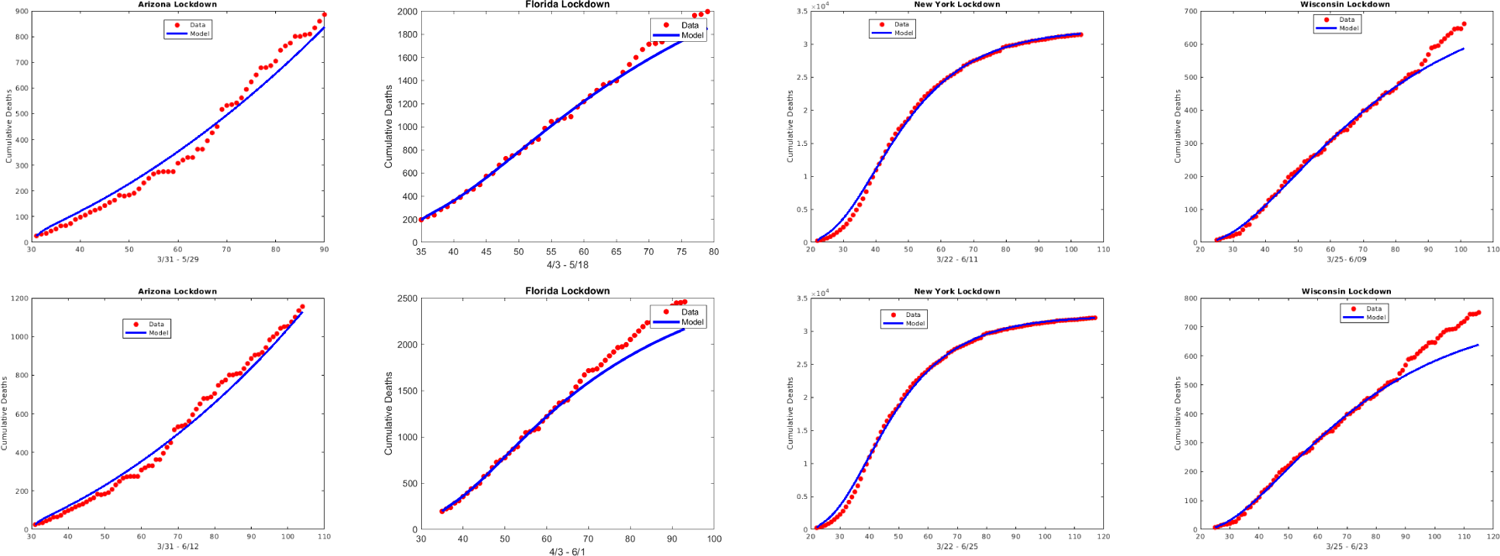
**Row 1**: The cumulative deaths for each state if lockdown was lifted 2 weeks later. **Row 2**: The cumulative deaths for each state if lockdown was lifted 4 weeks later.

These varying lockdown dates scenarios can be used to assess the efficacy of lockdown periods and to guide the manner in which the lockdown periods are implemented. For example, one interesting lockdown scenario is to consider what would happen if the entire U.S. implemented a lockdown during the month of December 2020 for two weeks. In order to simulate such a strategy we reduce the contact rate parameters - i.e., the betas - by 40% [11] and run the simulation forward. As shown in Figure 7 below, the simulations give that such a lockdown would result in a marked decrease in the number of deaths.

**Figure 7:**
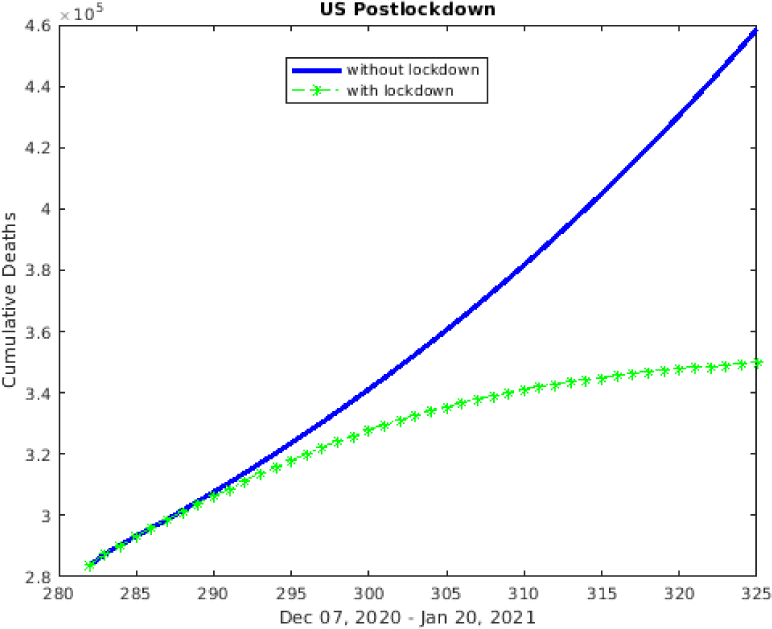
Cumulative deaths in US with lockdown in December (green) versus without lockdown (blue)

**Figure 8:**
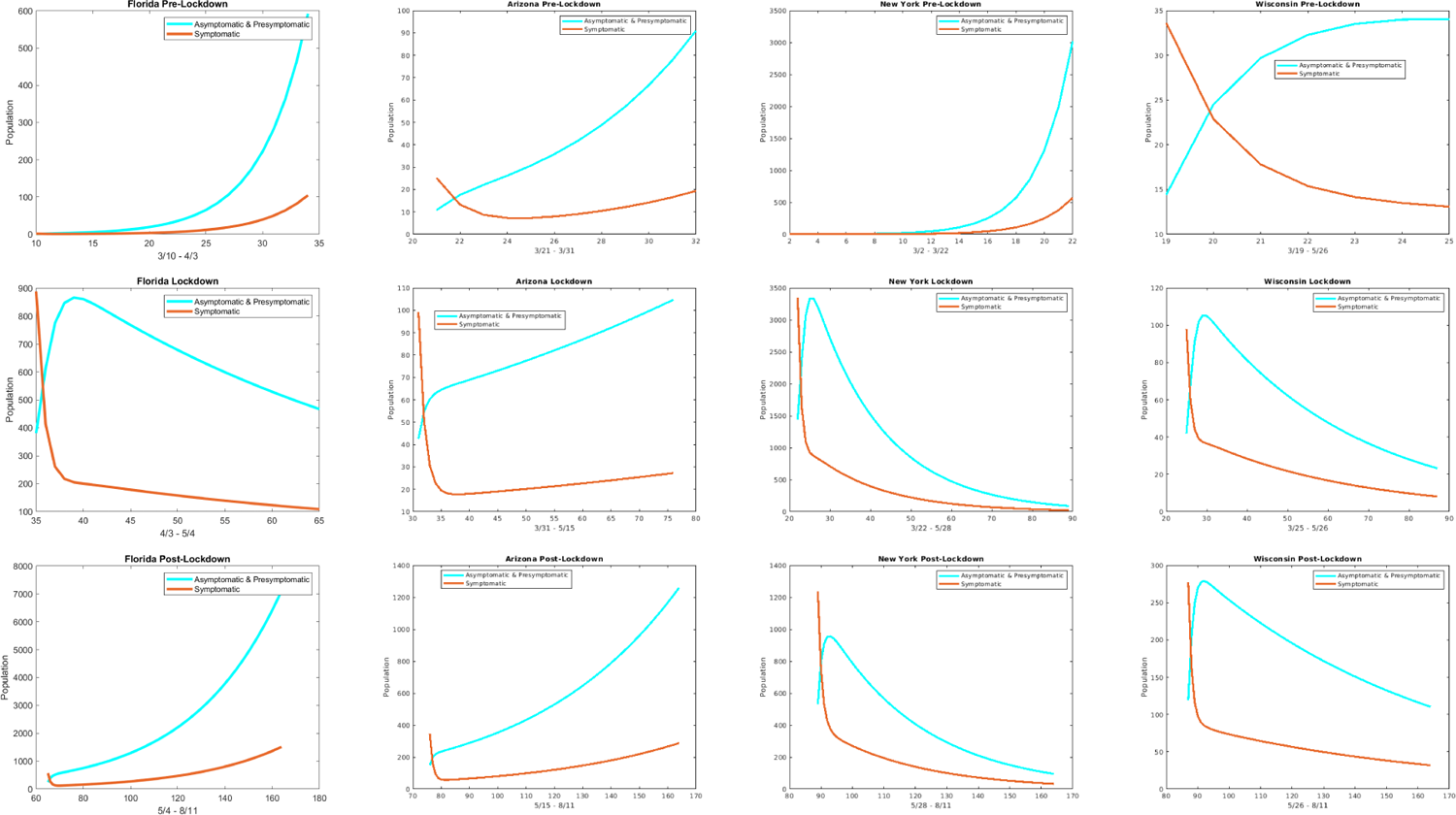
The plots show the pre-symptomatic and asymptomatic versus symptomatic individuals for each state during their respective time period. **Row 1**: Pre-lockdown period. **Row 2**: Lockdown period. **Row 3**: Post-lockdown period.

#### 4.3.2. Simulations for Asymptomatic, pre-symptomatic, and Symptomatic

In this section, we focus more on comparing the contributions of the symptomatic infectious group versus the asymptomatic and pre-symptomatic infectious group. To this end, we first consider plotting the daily cases for the two groups against each other. In all states and time periods, we find that the number of daily cases for the asymptomatic and pre-symptomatic is more than the daily cases for the symptomatic as shown in the following figure.

The parameter *β_I_* represents the contacts by the symptomatically infectious individuals that could transmit the disease. By setting *β_I_* = 0, we eliminate that transmission and focus on the effect of the pre-symptomatic and asymptomatic individuals. Similarly, fixing *β_A_* = *β_P_* = 0, targets the effect of the symptomatically infectious. In order to further discern the role of the asymptomatic and pre-symptomatic infectious versus that of the symptomatic infectious, we consider these zero contact rate scenarios in terms of the number of daily cases for all of the states and the entire US. In Figure 9, we show the results for the pre-lockdown and post-lockdown periods and for those periods, we again get that the larger contribution comes from the asymptomatic and pre-symptomatic individuals. For the lockdown period we obtain mixed results and these are considered in the appendix (Figure 16).

**Figure 9:**
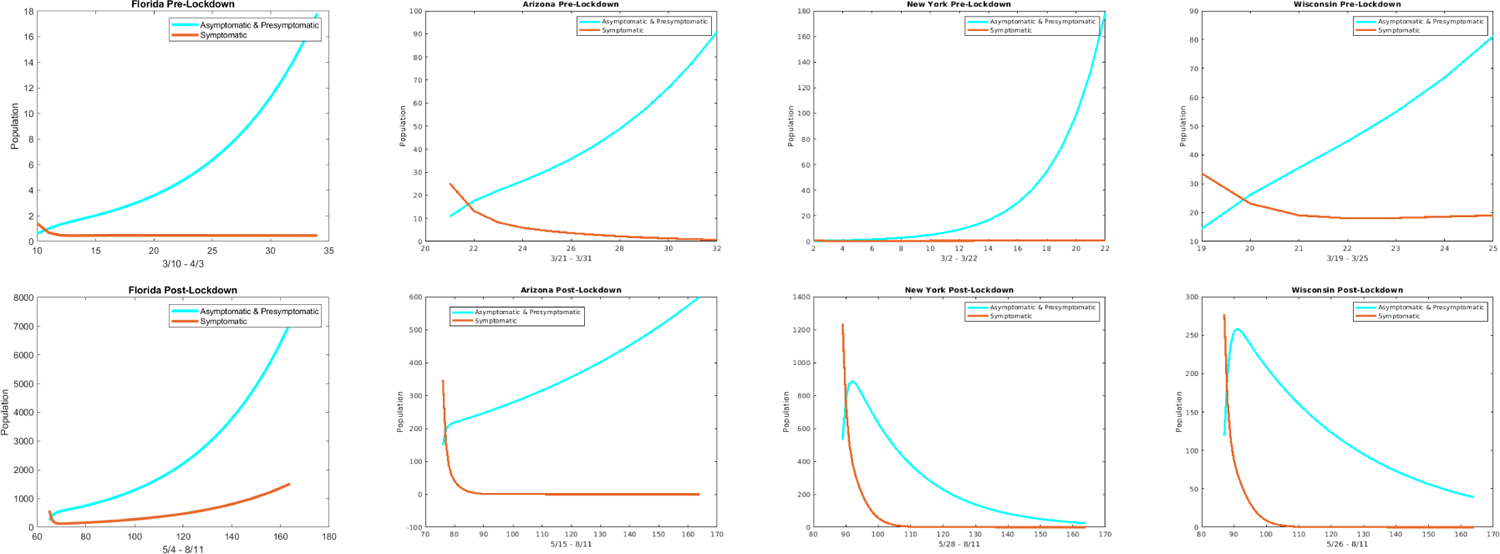
The plots show the daily cases for each state with *β_I_* = 0 (cyan curve) and with *β_A_* = *β_P_* = 0 (red curve) during their respective **Row 1**: Pre-lockdown period and **Row 2**: Post-lockdown period.

In Figure 10, we again take the *β_I_* = 0 (green curve) versus the *β_A_* = *β_P_* = 0 (magenta curve) scenario, but here we consider the effect in terms of cumulative cases and cumulative deaths for the entire US and the state of Arizona. Both types of plots indicate that the main drivers are the asymptomatic and pre-symptomatic individuals.

**Figure 10:**
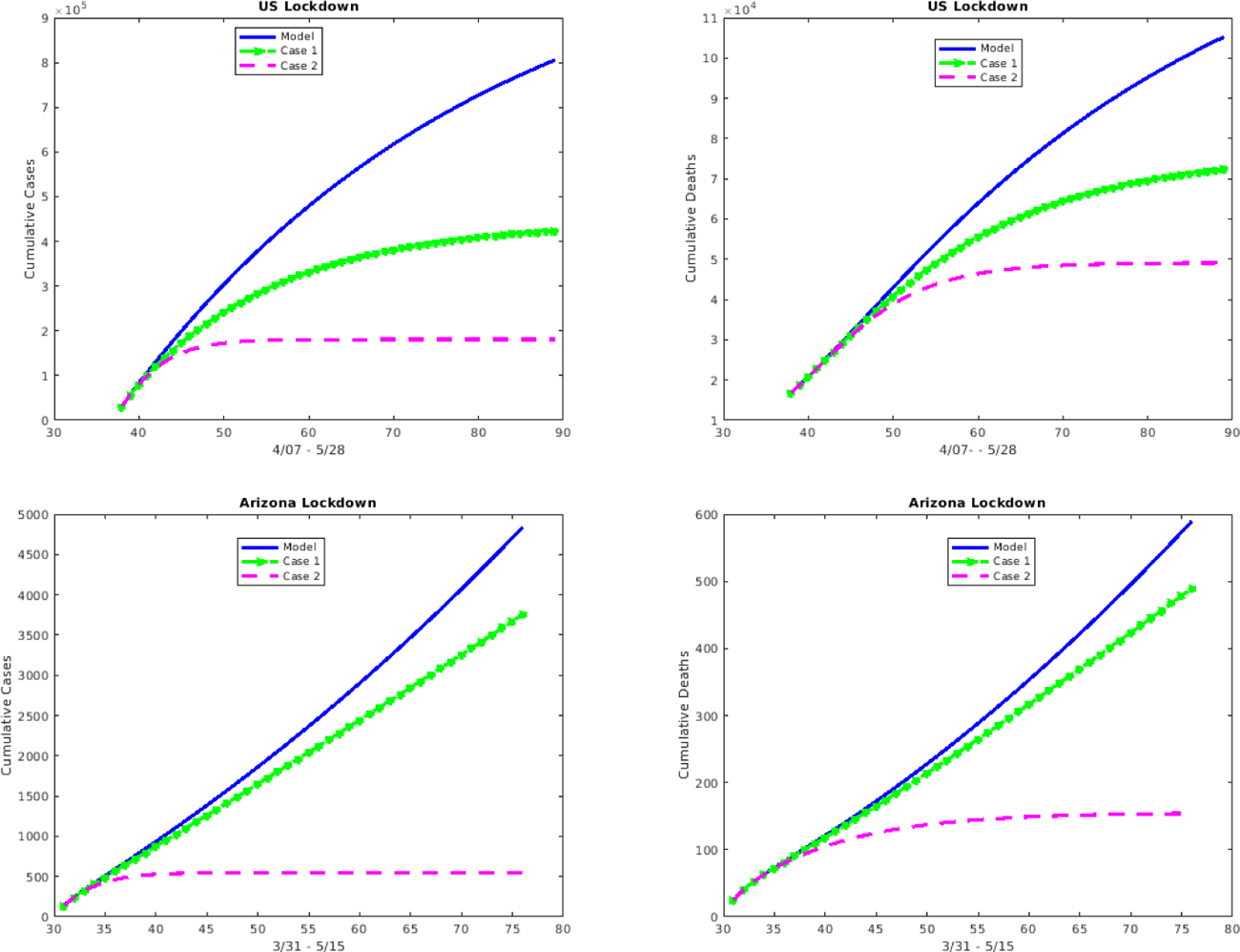
The cumulative cases and deaths for the US and Arizona over the respective lockdown period where **Case 1** corresponds to *β_I_* = 0 (green curve; so focuses on the impact of asymptomatic/pre-symptomatic individuals) and **Case 2** corresponds to *β_A_* = *β_P_* = 0 (magenta curve; so focuses on the impact of symptomatic individuals)

#### 4.3.3. Isolating Symptomatic and Asymptomatic/Pre-symptomatic

Recall that the *τ* values give the rate for testing/detection of infected individuals and subsequent self-isolation. In this section we compare and contrast the effect of isolating the symptomatic against the effect of isolating the asymptomatic and pre-symptomatic by changing the appropriate *τ* values. For example, to address the effect of isolating symptomatic infectious individuals, we can increase the value of *τ_I_* (the rate that symptomatically infectious individuals self-isolate) while keeping all other rates the same. We consider this scenario for the entire US and find that when *τ_I_* increases (so that the number of symptomatically infectious individuals that are self-isolated and taken out of the general population increases), the number of cumulative deaths decreases by as much as 12.5%. Alternatively, if the rate of self-isolation for the asymptomatic and pre-symptomatic infectious individuals - *τ_A_* and *τ_P_* respectively - are increased and the *τ_I_* is kept at the baseline value for the US, the number of cumulative deaths decreases by about 35% at the end of the period - see Figure 11. Thus, the decrease is more pronounced when the *τ_A_* and *τ_P_* are increased than when the *τ_I_* is increased which indicates that the asymptomatic and pre-symptomatic have more impact. Figure 12 focuses on the difference in impact with 50% increase in testing and shows that the asmptomatic and pre-symptomatic testing gives up to about four times as much percent decrease as the symptomatic.

**Figure 11:**
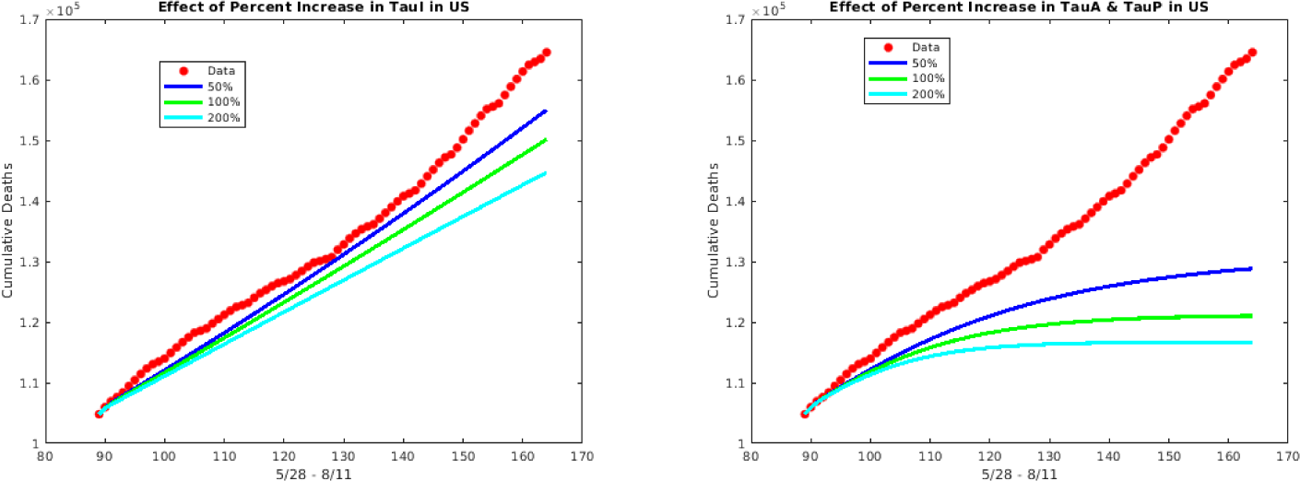
The effect of an increase in testing of symptomatic individuals versus asymptomatic and pre-symptomatic individuals on the cumulative deaths in the US during post-lockdown period 5/15 to 8/11.

**Figure 12:**
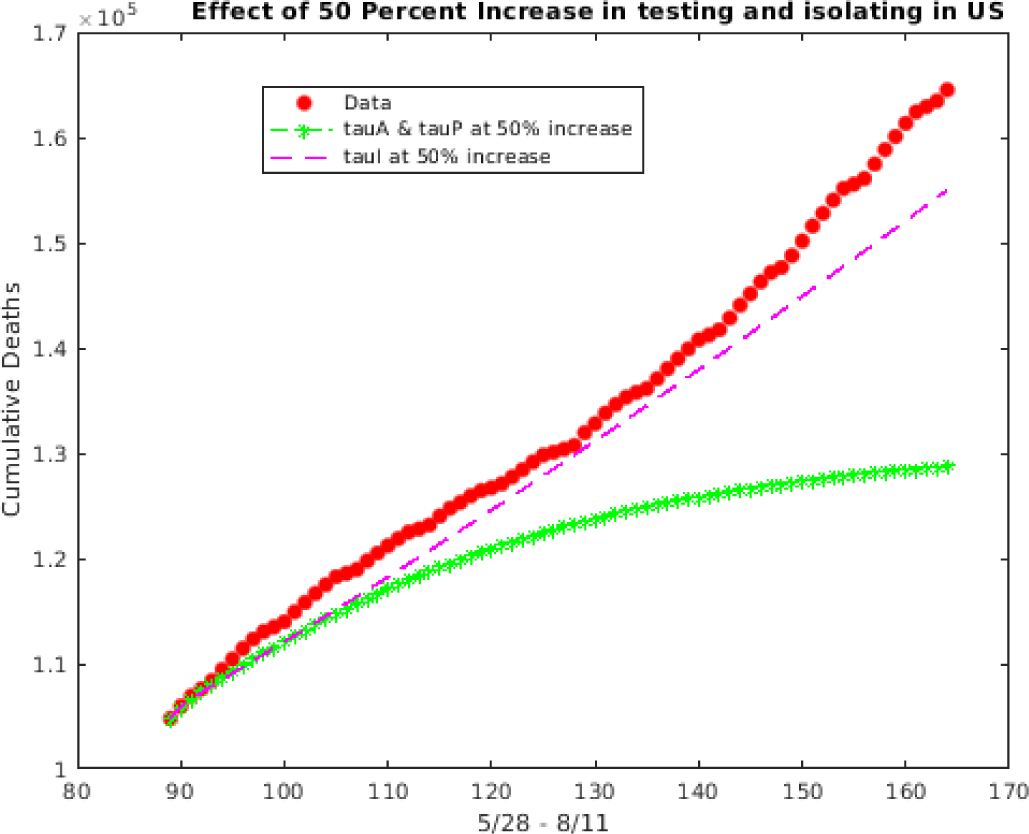
The effect of a 50% increase in testing of asymptomatic and pre-symptomatic versus symptomatic on the cumulative deaths in the US during post-lockdown period 5/28 to 8/11.

We can also compare the effect of these type of testing scenarios for different states. For example, although Wisconsin and Florida had similar testing rates around 12/10/2020, we find that a 50% increase in *τ_A_* and *τ_P_*, results in roughly a 20% decrease in cumulative deaths for Wisconsin compared to a 50% decrease in cumulative deaths for Florida - Figure 13. Note that the results for both states show the greater influence of the aysmptomatic and pre-symptomatic infectious.

**Figure 13:**
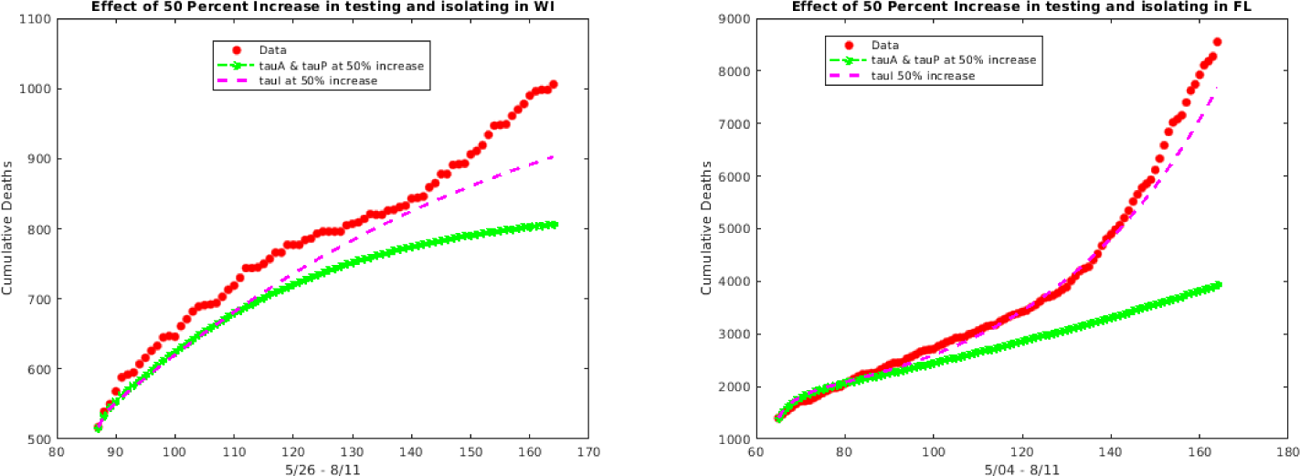
The effect of a 50 percent increase in testing of asymptomatic and pre-symptomatic (green curve) versus symptomatic (magenta curve) individuals on the cumulative deaths for Wisconsin and Florida.

#### 4.3.4. Minimum testing needed

In this section we consider the question concerning the minimum rate of testing/detection required to lower the control reproduction number, *R_C_* below one. This question is more difficult to answer directly, but we can begin to get at an answer by again considering the effect of increasing testing. For example, for the entire US from May 28 to Aug 11 (after lifting of lockdown) if the testing rate for asymptomatic and pre-symptomatic is 0.2 - that is, if *τ_A_* and *τ_P_* are both set to 0.2, while *τ_I_* = 0.4, then the reproduction control number *R_C_* is 1.0346 (so just above 1). If *τ_A_* and *τ_P_* are bumped above 0.2, for example to 0.25, while *τ_I_* remains at 0.4, then *R_C_* falls below 1 - to 0.95178 - and the resulting decrease in cumulative deaths and daily cases is shown in Figure 14. To get a better appreciation of the impact of testing for asymptomatic and pre-symptomatic cases, note that if *τ_A_* and *τ_P_* are kept at their baseline fitted values, then *τ_I_* must be increased to 1.7 to get *R_C_* = 0.9945. Of course, the challenge with all of these testing scenarios is that the testing of asymptomatic and pre-symptomatic individuals is more difficult since those individuals do not have symptoms, so in order to achieve these testing rates, widespread and frequent testing should be implemented.

**Figure 14:**
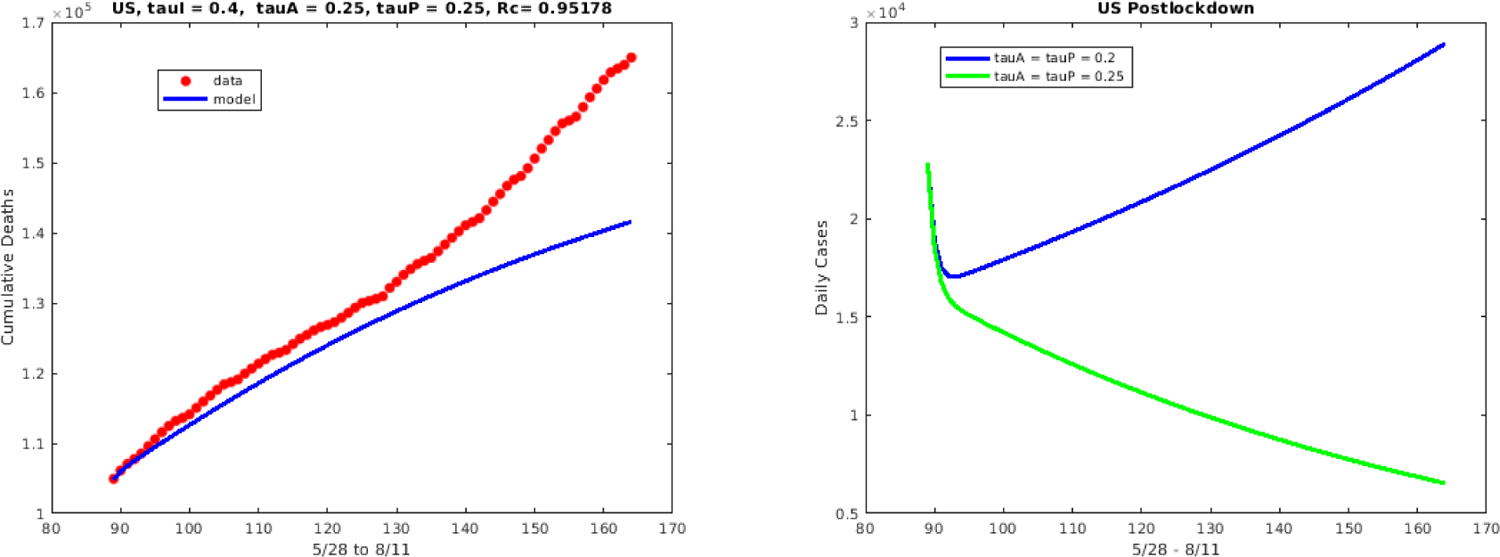
The effect of an increase in *τ_A_* and *τ_P_*

### 4.4. Forward Simulation of the Pandemic

Natural herd immunity refers to the process by which enough of the population achieves immunity via natural recovery from the disease, so that the remaining population that is not immune also receives protection against the acquisition of the disease. The process involves a natural herd immunity threshold, that is, the minimum number of people required to achieve disease-acquired immunity. For a disease with a basic reproduction number of *R*_0_, the necessary minimum fraction of people who must achieve immunity can be given as 1 *−* 1*/R*_0_. We simulate for both Wisconsin (a state which was considered an epicenter later in 2020) and the entire US, a scenario in which COVID-19 is allowed to run its natural course (with no implementation of any new control or mitigation strategies) until natural herd immunity is achieved. For both Wisconsin and the entire US, the number of deaths that occur before reaching the herd immunity threshold, is considerably high; thus, making the success of a natural herd immunity strategy for COVID-19 questionable (at the very least). For the state of Wisconsin, the number of deaths that occurs is approximately 80,000(Fig 15A)and for the entire U.S., the number of deaths is more than 5 million (Fig 15B).

**Figure 15:**
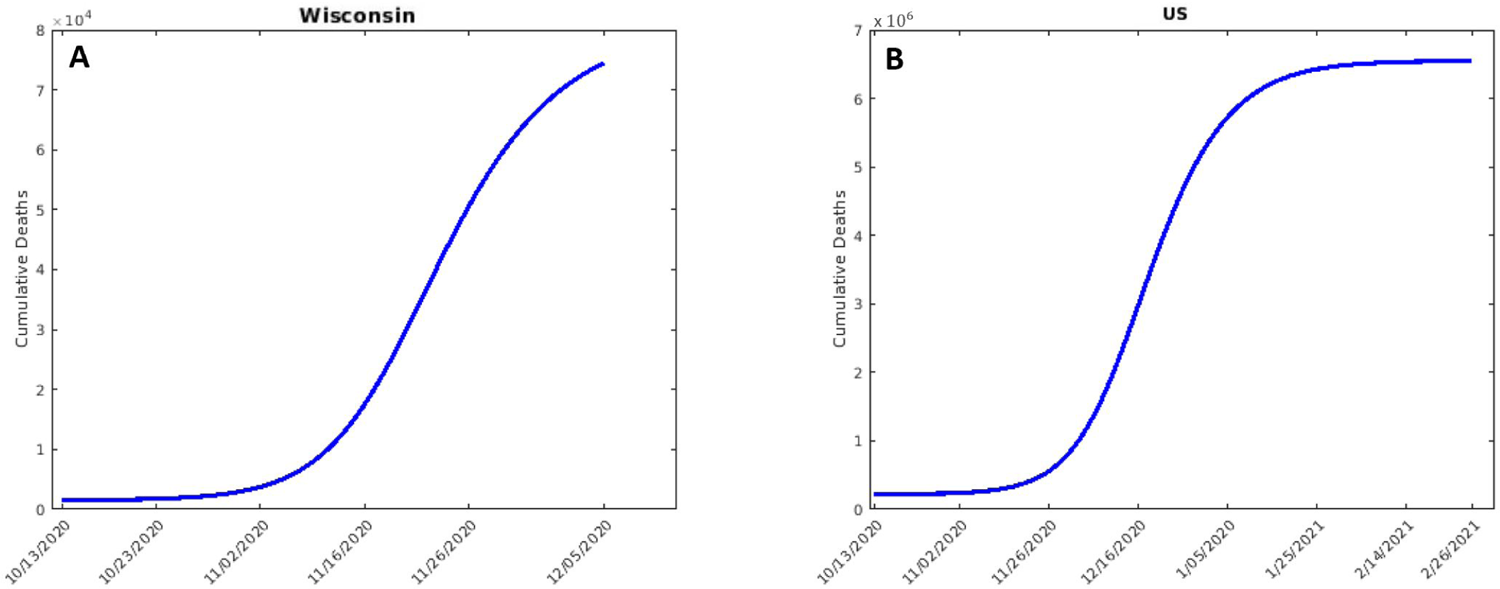
The number of deaths that will occur before natural herd immunity threshold is achieved for the state of Wisconsin (A) and the entire US (B). The simulations are run starting on October 13.

## 5. Summary

From various perspectives, our analysis indicates that overall, the drivers of COVID-19 are the asymptomatic and pre-symptomatic infectious individuals. Even with vaccines, it is critical to control the spread. We see that if testing is increased, these asymptomatic and pre-symptomatic infectious have a greater impact than the symptomatic infectious; hence, widespread and frequent testing is key to controlling the spread of the disease. This is especially true after holiday periods and large gathering events which are often notorious for reduced and lax implementation of control and mitigation strategies such as social distancing. Thus, in the schools and workplaces, testing should be intentionally and systematically increased after such occurrences in order to control the silent transmission by the newly infectious individuals who do not (yet) have symptoms. Our simulations also show that although lockdown might be difficult in terms of economic impact, the benefit of even a short lockdown (2 weeks) in reducing deaths is considerable.

## Data Availability

Data is taken from the COVID-19 Data Repository by the Center for Systems Science and Engineering (CSSE) at Johns Hopkins University (2020)

## Acknowledgements

The authors extend special thanks to the 2020 African Diaspora Joint Mathematics Workshop (ADJOINT), Dr. Abba Gumel (our ADJOINT group leader) and the Mathematical Sciences Research Institute (MSRI) - especially the MSRI IT members Aaron Hale & Nate Orange. We also acknowledge the following funding sources for ADJOINT: Alfred P. Sloan Foundation (G-2020-12602), National Science Foundation (DMS-1915954 and DMS-2016406), and National Security Agency (H98230-20-1-0015).

## 7 Appendix A

### 7.1. Tables of State Variables and Parameter Values

**Table 3:** State variables for the Model 1

| State Variable | Description |
| --- | --- |
| $S$ | Population of susceptible individuals |
| $E$ | Population of non-quarantined exposed individuals (infected but not showing symptoms and cannot transmit infection; newly-infected but not infectious) |
| $E_p$ | Population of pre-symptomatic (infectious) individuals |
| $I$ | Population of symptomatically-infectious individuals |
| $A$ | Population of asymptotically-infectious individuals |
| $J$ | Population of self-isolated individuals |
| $H$ | Population of hospitalized individuals |
| $R$ | Population of recovered individuals |
| $C$ | Population of individuals in intensive care unit (ICU) |
| $D$ | Population of COVID-19 deceased individuals |

**Table 4:**
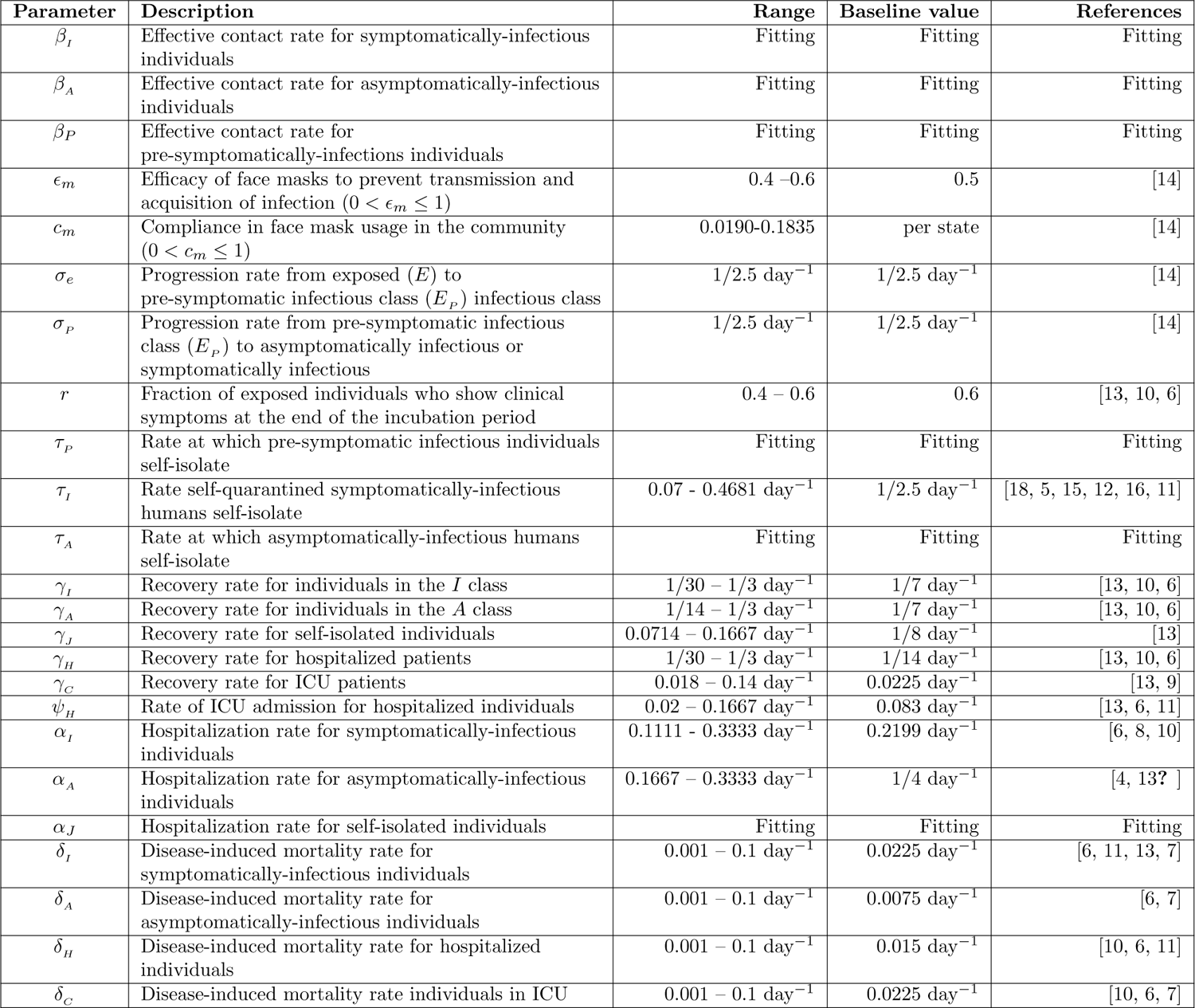
Parameter notation, description, values and sources for the Model 1. Some values change depending on the state and time period and for simulations that are post post-lockdown or involve testing implications, the testing parameter values are chosen to moreso align with reported testing trends.

#### 7.2. Lockdown period with separated symptomatic and asymptomatic individuals

In Figure 9, the results for the pre-lockdown and post-lockdown periods confirm the hypothesis that the asymptomatic and pre-symptomatic are the primary contributors to the spread of COVID-19. However, when we consider this scenario of targeting the asymptomatic and pre-symptomatic (by setting *β_I_* = 0) versus the symptomatic (with *β_A_* = *β_P_* = 0) for lockdown, we get mixed results. In particular for New York and Wisconsin we get more symptomatic infectious cases than asymptomatic and pre-symptomatic - see Figure 16.

**Figure 16:**
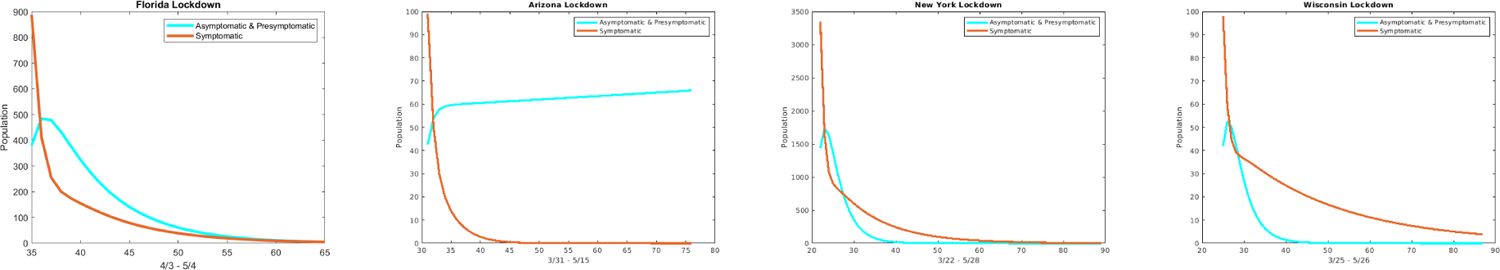
The plots show the daily cases for each state with *β_I_* = 0 (cyan curve) and *β_A_* = *β_P_* = 0 (red curve) for the lockdown period.

### 7.3. Goodness of Fit

In this section, we show the plots for the goodness of fit of the lockdown period for each state and the US as a whole.

**Figure 17:**
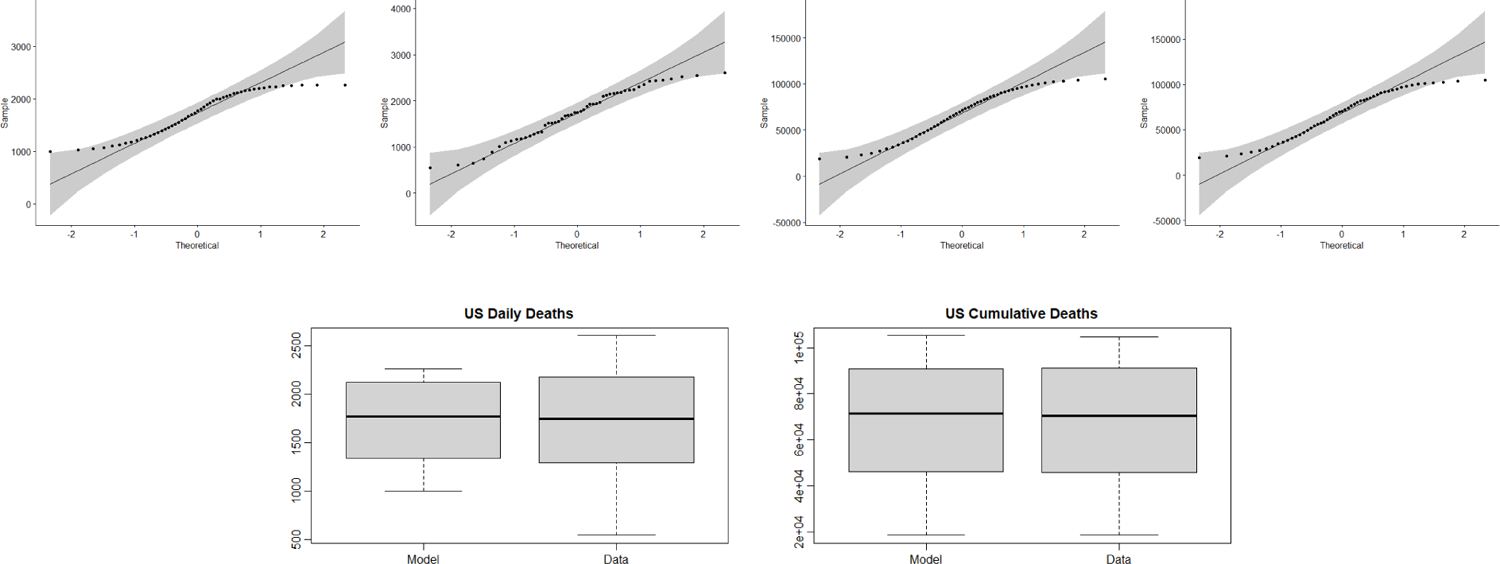
US **Row 1**: Q-Q plots for the (1) Daily death values from the model (2) Daily death values from the data (3) Cumulative death values from the model (4) Cumulative death values from the data. **Row 2**: Box plot of the (1) Daily death values from the model and data (2) Cumulative death values from the model and data.

**Figure 18:**
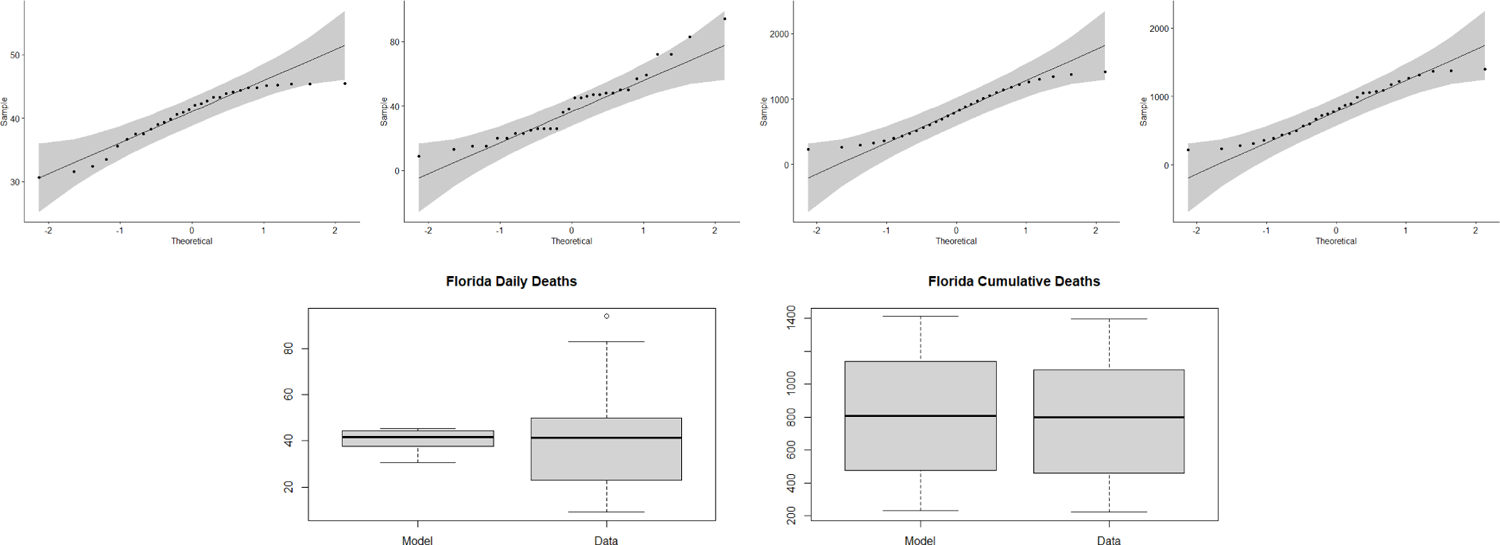
Florida **Row 1**: Q-Q plots for the (1) Daily death values from the model (2) Daily death values from the data (3) Cumulative death values from the model (4) Cumulative death values from the data. **Row 2**: Box plot of the (1) Daily death values from the model and data (2) Cumulative death values from the model and data.

**Figure 19:**
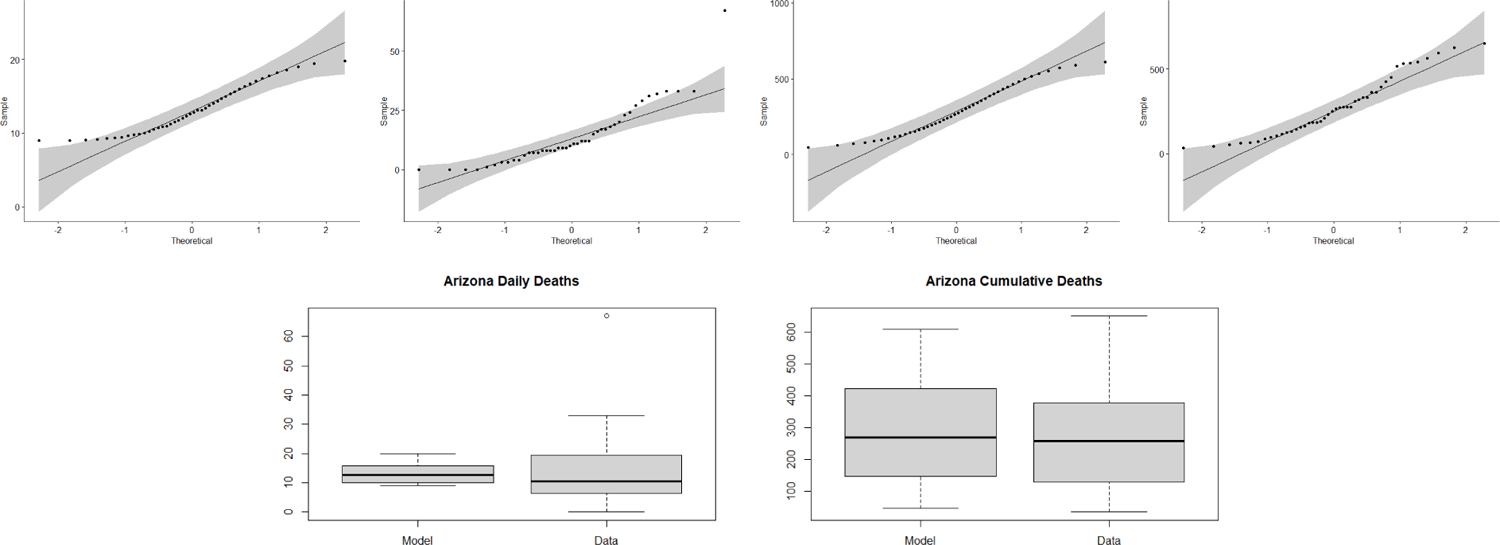
Arizona **Row 1**: Q-Q plots for the (1) Daily death values from the model (2) Daily death values from the data (3) Cumulative death values from the model (4) Cumulative death values from the data. **Row 2**: Box plot of the (1) Daily death values from the model and data (2) Cumulative death values from the model and data.

**Figure 20:**
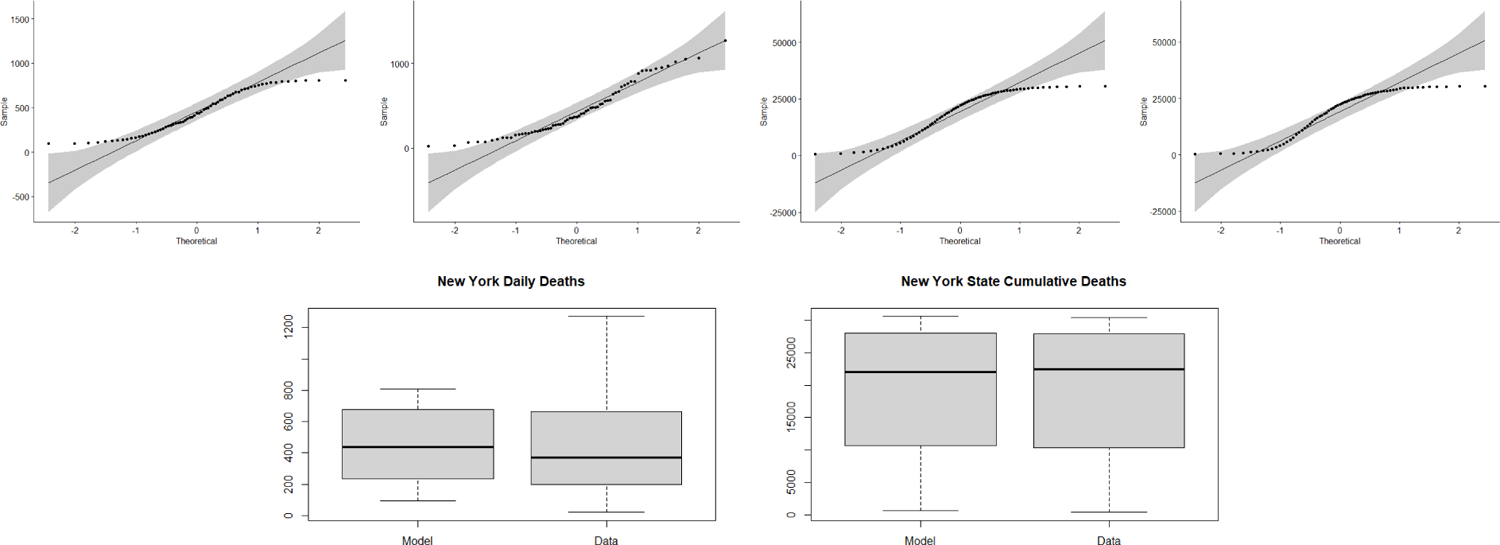
New York **Row 1**: Q-Q plots for the (1) Daily death values from the model (2) Daily death values from the data (3) Cumulative death values from the model (4) Cumulative death values from the data. **Row 2**: Box plot of the (1) Daily death values from the model and data (2) Cumulative death values from the model and data.

**Figure 21:**
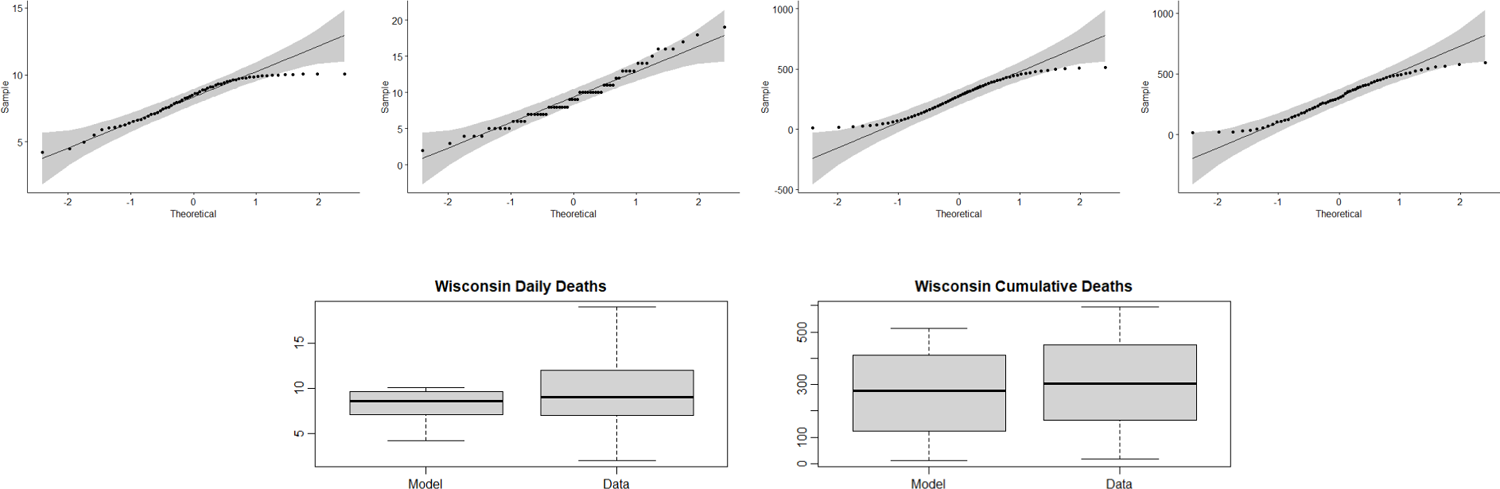
Wisconsin **Row 1**: Q-Q plots for the (1) Daily death values from the model (2) Daily death values from the data (3) Cumulative death values from the model (4) Cumulative death values from the data. **Row 2**: Box plot of the (1) Daily death values from the model and data (2) Cumulative death values from the model and data.

## Notes

### Competing Interest Statement

The authors have declared no competing interest.

### Funding Statement

ADJOINT at the Mathematical Sciences Research Institute

